# ACoRE: Accurate SARS-CoV-2 genome reconstruction for the characterization of intra-host and inter-host viral diversity in clinical samples and for the evaluation of re-infections

**DOI:** 10.1101/2021.01.22.21250285

**Authors:** Luca Marcolungo, Cristina Beltrami, Chiara Degli Esposti, Giulia Lopatriello, Chiara Piubelli, Antonio Mori, Elena Pomari, Michela Deiana, Salvatore Scarso, Zeno Bisoffi, Valentina Grosso, Emanuela Cosentino, Simone Maestri, Denise Lavezzari, Barbara Iadarola, Marta Paterno, Elena Segala, Barbara Giovannone, Martina Gallinaro, Marzia Rossato, Massimo Delledonne

## Abstract

We report Accurate SARS-CoV-2 genome Reconstruction (ACoRE), an amplicon-based viral genome sequencing workflow for the complete and accurate reconstruction of SARS-CoV-2 sequences from clinical samples, including suboptimal ones that would usually be excluded even if unique and irreplaceable. We demonstrated the utility of the approach by achieving complete genome reconstruction and the identification of false-positive variants in >170 clinical samples, thus avoiding the generation of inaccurate and/or incomplete sequences. Most importantly, ACoRE was crucial to identify the correct viral strain responsible of a relapse case, that would be otherwise mis-classified as a re-infection due to missing or incorrect variant identification by a standard workflow.

## BACKGROUND

The coronavirus disease 2019 (COVID-19) pandemic has thus far resulted in the infection of more than 84 million people, causing at least 1.8 million deaths (Johns Hopkins University, 1/1/2021)[1]. The agent responsible for COVID-19 is a β-coronavirus known as severe acute respiratory syndrome-associated coronavirus 2 (SARS-CoV-2) with a compact single-stranded RNA genome of 29,903 nucleotides. The first SARS-CoV-2 genome sequence was published soon after the initial outbreak [2], and more than 260,000 complete genome sequences have subsequently been deposited in the GISAID database [3]. The phylogenetic analysis of genomic sequences provides a valuable tool to track viral diversity during the course of a pandemic and to identify the emergence of prevalent strains characterized by lineage-specific single nucleotide variants (SNVs), such as the D614G variant in the SARS-CoV-2 spike protein gene (23403:A→G) [4–6]. As the virus propagates in human-to-human transmission, changes in the reference genome sequence must be recorded to monitor correlations between viral genotype and disease communicability, manifestation and severity [4,7–9]. The combination of genomic analysis and epidemiological data can also reliably determine the extent of SARS-CoV-2 transmission in different nations [10–12] and thus facilitates early decision-making to control local transmission [13]. Finally, mutations that may be relevant to the fitness or antigenic profile of the virus can be identified to ensure the efficacy of vaccines and immunotherapeutic interventions in the clinic[4,14].

Consensus variations reflect the analysis of virus sequences that differ between patients, but the analysis of intra-individual single nucleotide variations (iSNVs) is also important because it helps us to understand more about virus–host interactions, as previously demonstrated for Ebola, Zika, influenza and HIV [15–19]. The analysis of iSNVs during the COVID-19 pandemic may also provide data about the potential of SARS-CoV-2 for immunological escape and resistance to therapy, as well as on the sensitivity of molecular diagnostic assays [20–22]. However, the identification of iSNVs in clinical samples can be challenging because current protocols often feature enrichment and amplification steps that introduce technical errors indistinguishable from true biological variants [23].

The reconstruction of complete and accurate genomic sequences to detect both SNVs and iSNVs is therefore necessary to produce reliable data, at all these aims. In addition, the accumulation of meaningful data during pandemics requires the analysis of many samples, and the corresponding methods must therefore be cost-effective, straightforward and suitable for high-multiplexing [24]. The protocols must also be sensitive enough to detect low viral titers but applicable over a wide dynamic range of virus concentrations to allow the analysis of clinical samples with different viral loads, ideally including samples from early and late infection stages, that usually show a lower viral detection, or from re-infection/relapse cases [25,26].

Among the many approaches available for SARS-CoV-2 whole-genome analysis, the amplicon-based sequencing method developed by the ARTIC Network [27] is currently the most widely used [13,24,28–32]. Based on the PrimalSeq protocol originally developed for Zika virus [23,33], the ARTIC Network designed a set of 98 tiled amplicons in two PCR pools for the targeted whole-genome amplification of SARS-CoV-2 [27]. This approach is simple and highly sensitive, but it suffers from technical biases leading to uneven genome coverage, thus reducing the completeness and accuracy of genome sequencing, especially for the identification iSNVs in samples with low viral titers [34–36]. Sequencing technical replicates of multiple cDNAs generated from the same sample has been proposed as a mitigation strategy to identify iSNVs more reliably [23]. However, whereas amplicon-based sequencing has been widely used for the analysis of low-frequency variants [20–22,37,38] only a few studies thus far have evaluated the confidence of such calls and have implemented the sequencing of cDNA replicates to ensure accuracy [23]. False positives have also been reported among high-frequency variants supported by good sequencing depth, indicating that the risks of inaccurate sequencing are not limited to suboptimal samples [39].

To avoid the generation of incomplete genomic sequences typically associated with poor genome coverage [40–42], the sequencing of samples with fewer than 1000 virus copies per RT-qPCR reaction (Ct < 30) is currently discouraged [23,43]. However, the strict implementation of such recommendations would lead to the exclusion of many clinical samples, which are often unavoidably collected or stored under suboptimal conditions. Since specimens with these features may be unique and irreplaceable -central to the investigation conducted-, numerous studies therefore report sequencing data from samples with (very) low viral titers (Ct > 30) despite this advice [26,44,45]. To address these challenges, we set out to develop an optimized workflow, ACoRE (Accurate SARS-CoV-2 genome Reconstruction, for the reliable reconstruction of complete and accurate SARS-CoV-2 genomes from clinical samples with a broad range of Ct values, aiming to improve the flexibility, accuracy and throughout of amplicon-based sequencing.

## RESULTS

### Accuracy of SARS-CoV-2 genome reconstruction

The original Primalseq protocol stipulates two independent reverse transcriptions per sample and the subsequent amplification of the separate cDNAs in order to reduce technical errors. In this study, we initially tested replicate amplifications from the same cDNA to investigate whether this alternative approach could affect the reproducibility in the generation of SARS-CoV-2 consensus sequences and in the identification of intra-host variants. At this aim, we selected five COVID-19-positive swabs representing viral loads ranging from ∼500 to ∼2 million, based on Ct values (determined by RT-qPCR) ranging from 15.07 to 28.5 (**Table S1**). For each sample, we generated three cDNAs and carried out two separate amplifications, resulting in six replicates per starting RNA (**Figure 1A**). An individual KAPA library was prepared from each replicate, and sequencing in 250PE mode produced an average of 1 million fragments. The dataset was normalized to ∼800,000 fragments per library, corresponding to ∼7800× coverage per sample after alignment to the SARS-CoV-2 reference genome (**Table S3**).

**Figure 1.**
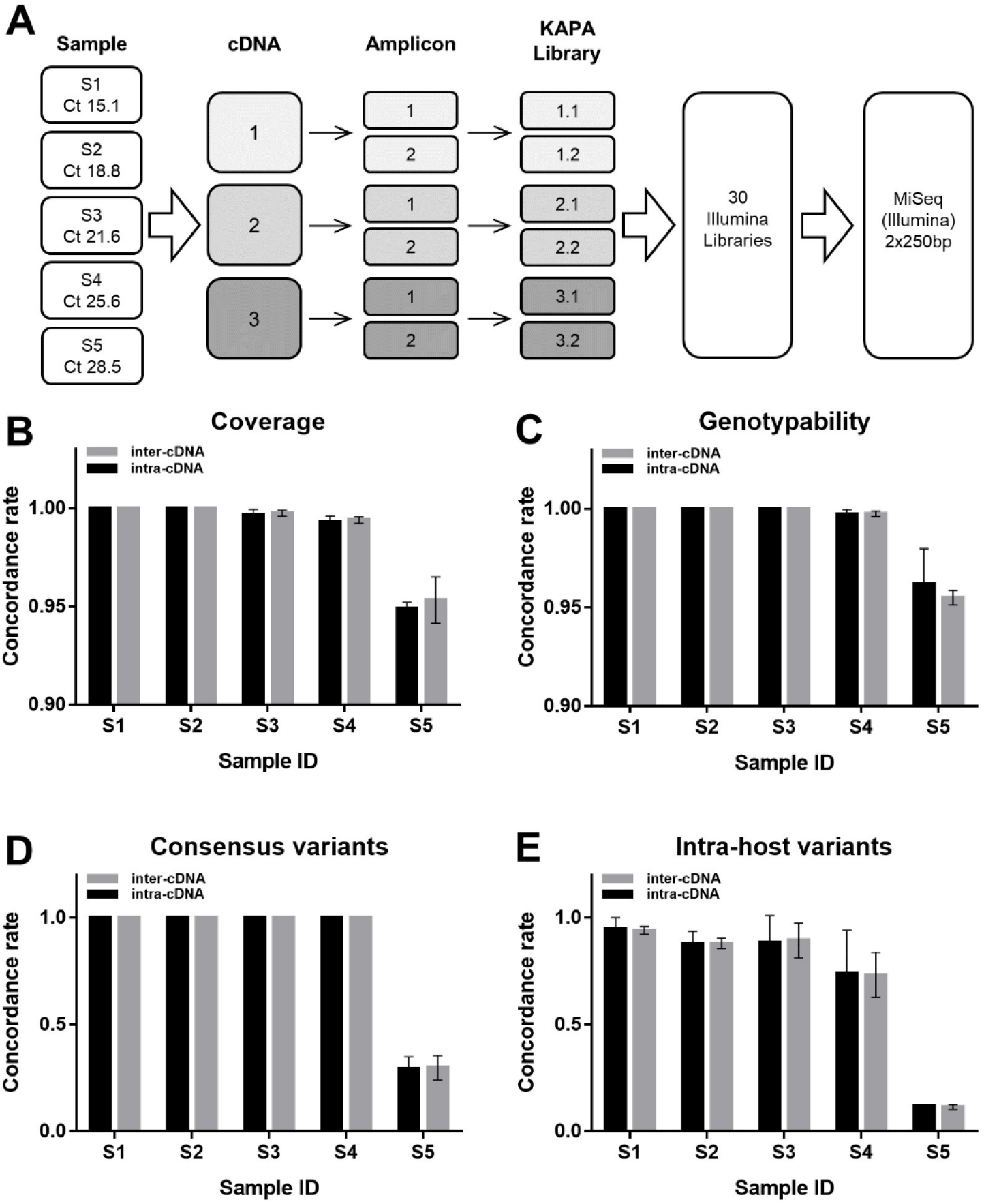
Comparison of intra-cDNA and inter-cDNA replicates of SARS-CoV-2 genome amplification and sequencing. **(A)** Schematic diagram showing the five clinical samples obtained from COVID-19 patients, their RT-qPCR Ct values and the experimental workflow. For each sample, we generated three independent cDNAs and each cDNA was amplified in duplicate using the ARTIC nCoV-2019 V3 Panel. Amplicons used as the input for library preparation were sequenced in 250PE mode on the Illumina MiSeq platform. The bar charts show mean concordance rates (± standard deviations) for **(B)** genome coverage, **(C)** genotypability, **(D)** consensus variants and **(E)** iSNV between amplification replicates generated from different cDNAs (inter-cDNA) or the same cDNA (intra-cDNA).

The sequencing coverage was variable across the different amplicons of the ARTIC panel, particularly in samples with a higher Ct value (**Figure 2 and Figure S1**). Interestingly, most amplicons showed either high (>500×) or very low (≤10×) to zero coverage, and amplicons absent in one replicate could be present in another, even when produced from the same cDNA. The concordance (R_c_) in sequencing coverage was high for replicates of four samples (R_c_ ∼0.99–1) but lower in sample S5 (R_c_ ∼0.95) with the lowest viral load (**Figure 1B and Table S4**), but there was no significant difference between replicates from the same or different cDNAs (p = 0.25, Wilcoxon test). Variations in coverage can affect genotyping accuracy, so we evaluated reproducibility in terms of genotypability by calculating the fraction of genomic positions where it is possible to call a genotype after aligning reads to the reference genome. The genotypability R_c_ was optimal or slightly lower than 1 in all samples (R_c_ = 0.99– 1), but lower in sample S5, which also showed the lowest sequencing coverage R_c_ (**Figure 1C and Table S5)**. Reproducibility was similar between inter-cDNA replicates and intra-cDNA replicates (p > 0.99, Wilcoxon test). To assess how fluctuations in genotypability and coverage affect the final viral genome sequences, we generated a consensus sequence for each replicate. The reproducibility among consensus variants was optimal in the first four samples, but consistently dropped to ∼0.3 for sample S5 (**Figure 1D and Table S6**). Nevertheless, reproducibility was again similar between inter-cDNA replicates and intra-cDNA replicates (p > 0.99, Wilcoxon test).

**Figure 2.**
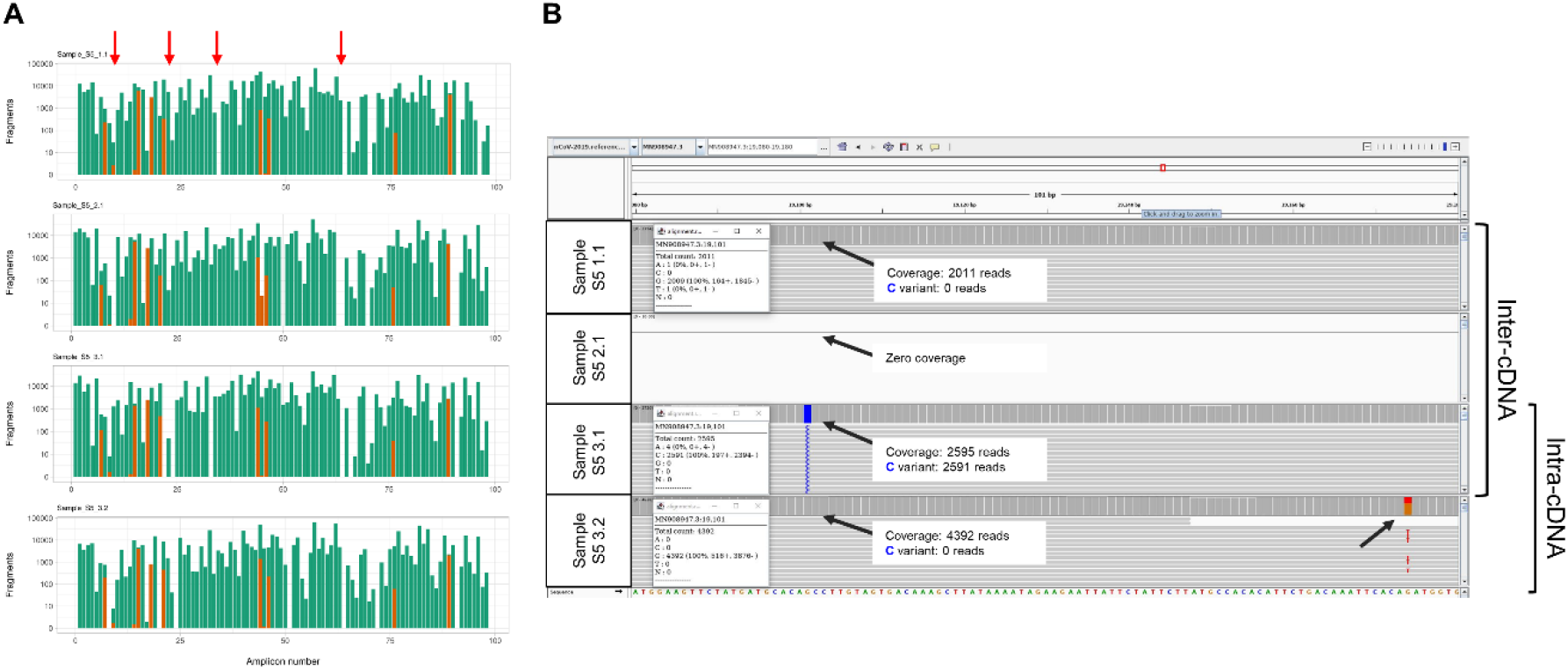
Coverage and variant calling between intra-cDNA and inter-cDNA replicates. **(A)** Sequencing coverage of the 98 amplicons of ARTIC V3 panel from four representative replicates of sample S5. Green bars represent the amplicons generated using the ARTIC original primer set, and orange bars represent the amplicons generated using the alternative V3 primers. Red arrows point at representative amplicons missing in only one replicate. **(B)** Integrative Genomics Viewer (IGV) visualization of four representative sequencing replicates of sample S5 in the region 19,080–19,180 of the SARS-Cov-19 genome. Black arrows indicate variants called only in one replicate. The amplicon was not amplified in replicate S5 2.1.

The number of iSNVs (frequency >3%) varied significantly between technical replicates, with a small fraction of iSNVs shared by different replicates compared to the total number of iSNVs identified (**Table S7**). The R_c_ was suboptimal (<0.95) for all samples and steadily decreased as the Ct value increased (**Figure 1E and Table S8**), but there was no significant difference between replicates generated from the same or different cDNAs (p = 0.44, Wilcoxon test). In summary, consensus sequences and intra-host variants can be strongly affected by uneven amplicon representation and PCR errors (**Figure 2**) confirming the need to sequence at least two replicates to achieve an accurate characterization of the SARS-CoV-2 genome. However, the two amplifications can be generated from the same starting cDNA, thus reducing sample consumption and costs.

### Improvement of genome reconstruction by merging technical replicates

While addressing the reproducibility issues observed for both SNVs and iSNVs in samples with low viral loads, we also tested whether merging two or more technical replicates could improve coverage and genotypability. The rationale was the observation that amplicons with the lowest coverage varied across different replicates, and amplicons missing in one replicate could have a coverage >100× or >1000× in others (**Figure S1**). All possible combinations of two replicates for each sample were merged and downsampled to 800,000 fragments (400,000 for each replicate) to obtain the same sequencing input data as the initial analysis based on a single replicate (**Table S9**). When considering the merged datasets rather than single-replicate data, the average coverage consistently increased in the sample with the highest Ct value (p < 0.0001, Mann Whitney U-test), confirming that merging two amplification replicates (intra-cDNA or inter-cDNA) could mitigate the technical variability in amplicon coverage (**Figure 3A-C**) as well as significantly (p < 0.0001, Mann Whitney U-test) enhance the genotypability (**Figure 3B**). Merging up to six replicates achieved a slight further improvement in both coverage and genotypability (**Figure 3A-B**), indicating that both properties can be maximized by analyzing replicates of samples with low viral loads. Indeed, merging all sequence data available for sample S5 (with the lowest reproducibility) increased coverage sufficiently to achieve >96.98% non-ambiguous bases in the consensus sequence (**Figure 3C-D**), which is the GISAID threshold for classifying a SARS-CoV-2 genome as complete [3]. Similar improvement was achieved in a panel of 170 clinical samples analyzed in duplicate or quadruplicate (**Figure 3E-G** shows three representative samples).

**Figure 3.**
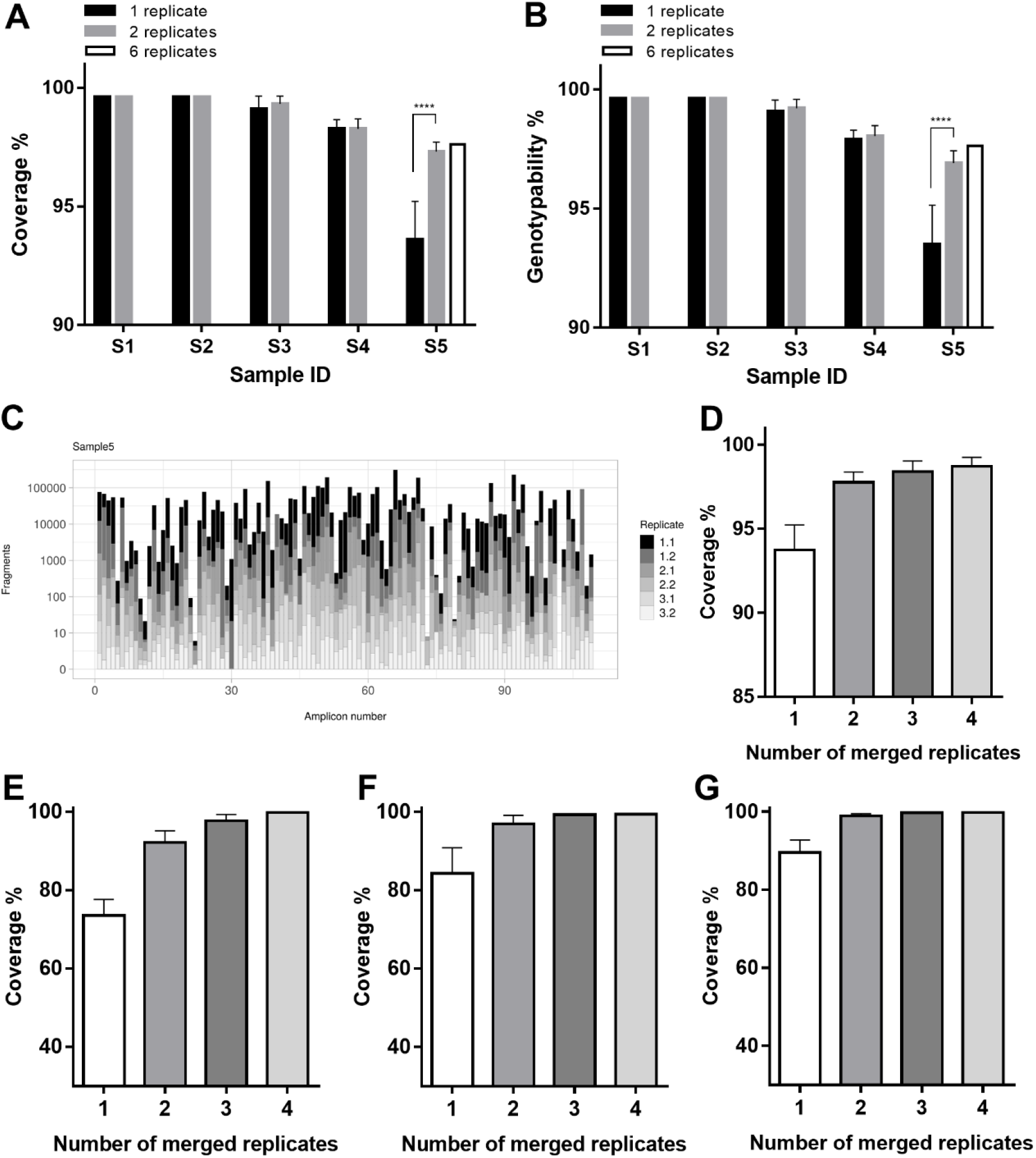
Merging sequencing replicates can improve coverage and genotypability. **(A)** Mean percentage genome coverage (± standard deviations). **(B)** Mean percentage genotypability (± standard deviations). Both genome coverage and genotypability were calculated for single replicates or after merging all possible combinations of two or six replicates, starting from the same total sequencing reads (****p < 0.0001, Mann Whitney U-test). **(C)** The coverage fraction contributed by each of the six replicates generated from sample S5. **(D)** Percentage of genome coverage after merging different numbers of replicates from sample S5, and from three other COVID-19-positive swab samples, namely samples 3270 **(E)**, 4572 **(F)**, 4173 **(E)**, whose sequencing results are reported in **Table S12**.

### Improvement of the technical workflow for viral genome sequencing

One drawback of the ARTIC protocol on the Illumina platform is the need for 250PE sequencing to cover the full length of the amplicons (400 bp). This type of sequencing is currently available only for MiSeq and NovaSeq6000 SP flow cells, increasing the cost per sample and reducing the sample throughput. We therefore generated shorter libraries using the NexteraFlex approach and tested the use of alterative flow cells (NextSeq500/550 and NovaSeq6000 S1) and sequencing mode (150PE) on the 30 samples originally tested using the KAPA library (**Figure 1A**). Despite skipping the laborious input DNA and library quantification steps before sequencing, the variability in the number of fragments analyzed per sample was lower (CV = 22.5%) than the full-amplicon approach (CV = 38.3%) described above (**Figure 4A**). The sequencing data were mapped to the reference genome (**Table S10**) and compared to the 250PE dataset (KAPA library) normalized with the same average-mapped coverage as the 150PE dataset (NexteraFlex library) (**Table S11**). Sequencing coverage was evenly distributed along the amplicons even when the NexteraFlex protocol was used, because the partial overlap of ARTIC amplicons compensated for the expected loss of sequence representation at the amplicon ends due to tagmentation (**Figure 4B**). The sequencing of fragmented amplicons had no adverse impact on genome coverage and genotypability, which were significantly higher compared to the full-length amplicon sequencing (p < 0.001 and p = 0.024, respectively, Friedman test; **Figure 4C-D**). Despite the lower coverage, similar results were observed with 100PE sequencing simulated after trimming the 150PE dataset (**Figure 4C-D**). The fragmented-amplicon approach was therefore advantageous for multiple aspects of SARS-CoV-2 sequencing, by increasing coverage, genotypability and throughput (allowing higher multiplexing) while reducing sequencing costs and eliminating unnecessary protocol steps such as DNA quantification after PCR and library quantification before pooling.

**Figure 4.**
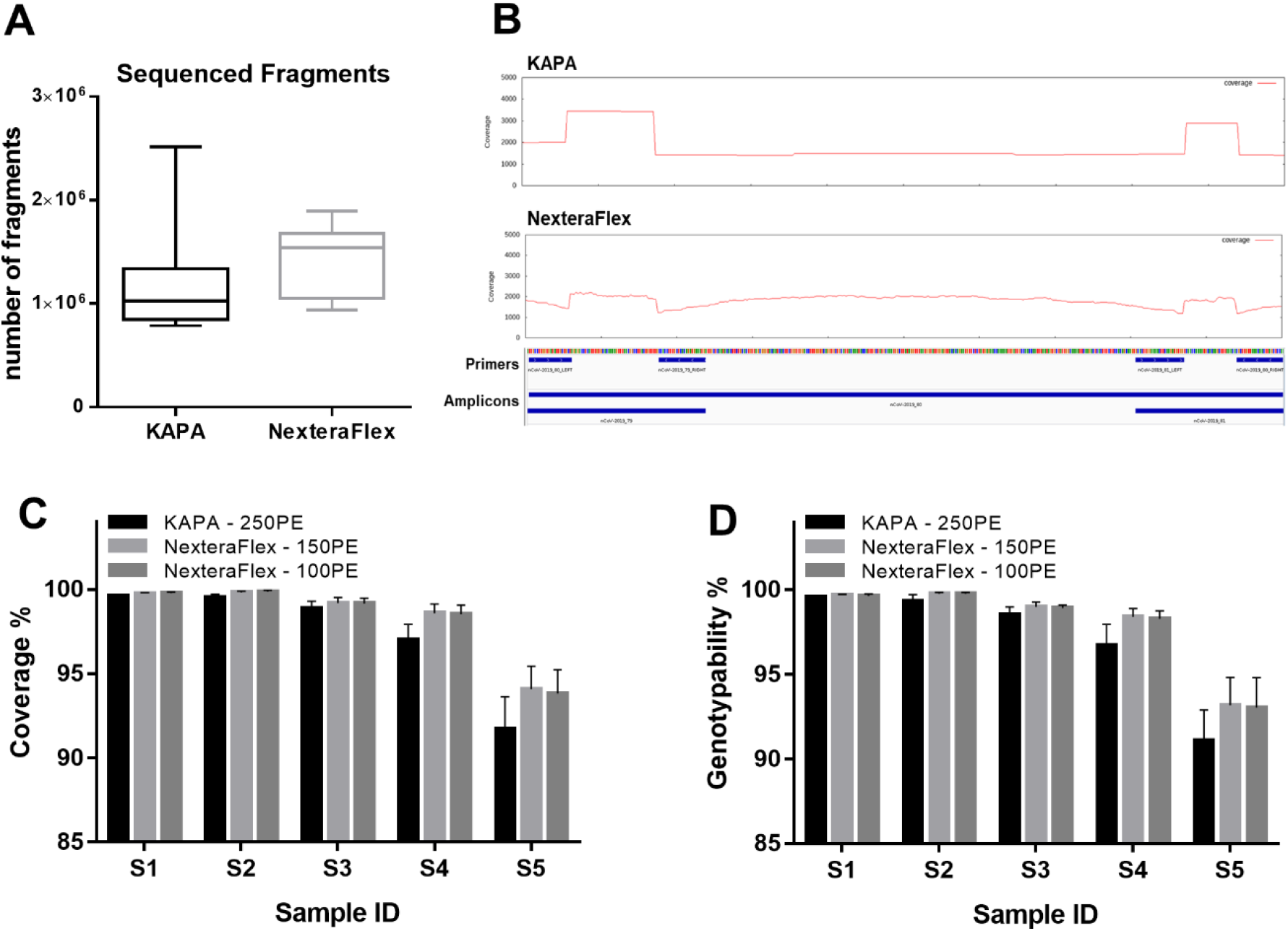
Comparison of SARS-CoV-2 sequencing and mapping results obtained using the KAPA and NexteraFlex library preparation kits. **(A)** Distribution of the number of fragments generated using the KAPA and NexteraFlex kits for the same set of 30 replicates. **(B)** Visualization of mean sequencing coverage on a representative ARTIC amplicon using the KAPA and NexteraFlex library kits. Given the overlap with adjacent amplicons, the 5′ and 3′ ends show increased coverage. **(C)** Mean coverage (± standard deviations) and **(D)** mean genotypability (± standard deviations) of sequencing libraries prepared from the 30 replicates using either the KAPA or NexteraFlex kits. The 100PE results were obtained from the 150PE dataset by *in silico* trimming.

Although the NexteraFlex protocol saves on costs, this is offset by the requirement for multiple sequencing replicates from the same sample to improve genome coverage. We therefore compared the effect of sequencing a library generated from two replicates (each amplified from 5 µL of cDNA) and a standard library prepared from a single amplification generated from double amount of cDNA (10 µL). Because samples with a low viral load benefit the most from multiple replicates, we analyzed 20 samples with a Ct range of 25–35 (**Figure S2A**). Two samples showed a lower coverage in libraries produced from a single cDNA, but overall there was little difference in coverage (p = 0.1) or genotypability (p = 0.09) when comparing the two conditions (Wilcoxon test; **Figure S2B-C**). This result confirmed that the reconstruction of SARS-CoV-2 genomes can also be maximized by increasing the amount of template cDNA through the use of more complex samples. Although such adjustments can improve coverage and genotypability, technical replicates are still required for the identification of true-positive variants.

### Application of the optimized workflow to large sets of samples

Next we applied the optimized workflow to a set of 170 clinical samples representing a wide range of viral loads, with Ct values in the range 15–40 (**Figure S3**). Each sample was amplified in duplicate or quadruplicate starting from 10 µL cDNA, and 100PE sequencing was carried on a NovaSeq6000 SP flow cell using NexteraFlex libraries, generating an average of ∼2.8 million fragments per replicate (**Table S12**). After pooling data from the replicates, ∼75% of the samples showed both coverage and genotypability >96.98% (**Figure 5A-B**) which is a clear improvement over the sequencing of a single cDNA (**Figure 5C-D**). Most (90.9%) of the samples that were not fully reconstructed were characterized by a low viral load (Ct > 30), but almost half (45%) of the samples in this Ct value range were nevertheless reconstructed optimally (**Figure 5E-F**). In particular, five of the seven viral genomes from swabs with a Ct value ≥ 38 were completely reconstructed (>96.98%), indicating that the outcome is not solely determined by the viral titer in the starting material. In order to generate accurate consensus sequences, we applied the same approach used to identify true-positive iSNVs (only variants in both replicates were included in the final consensus). This approach revealed that 22 samples (12.94%), with Ct 25.9-40, would have included at least one false-positive variant in the consensus sequences based on single-cDNA analysis, but these were efficiently removed by considering the concordance between replicates (**Table S13**).

**Figure 5.**
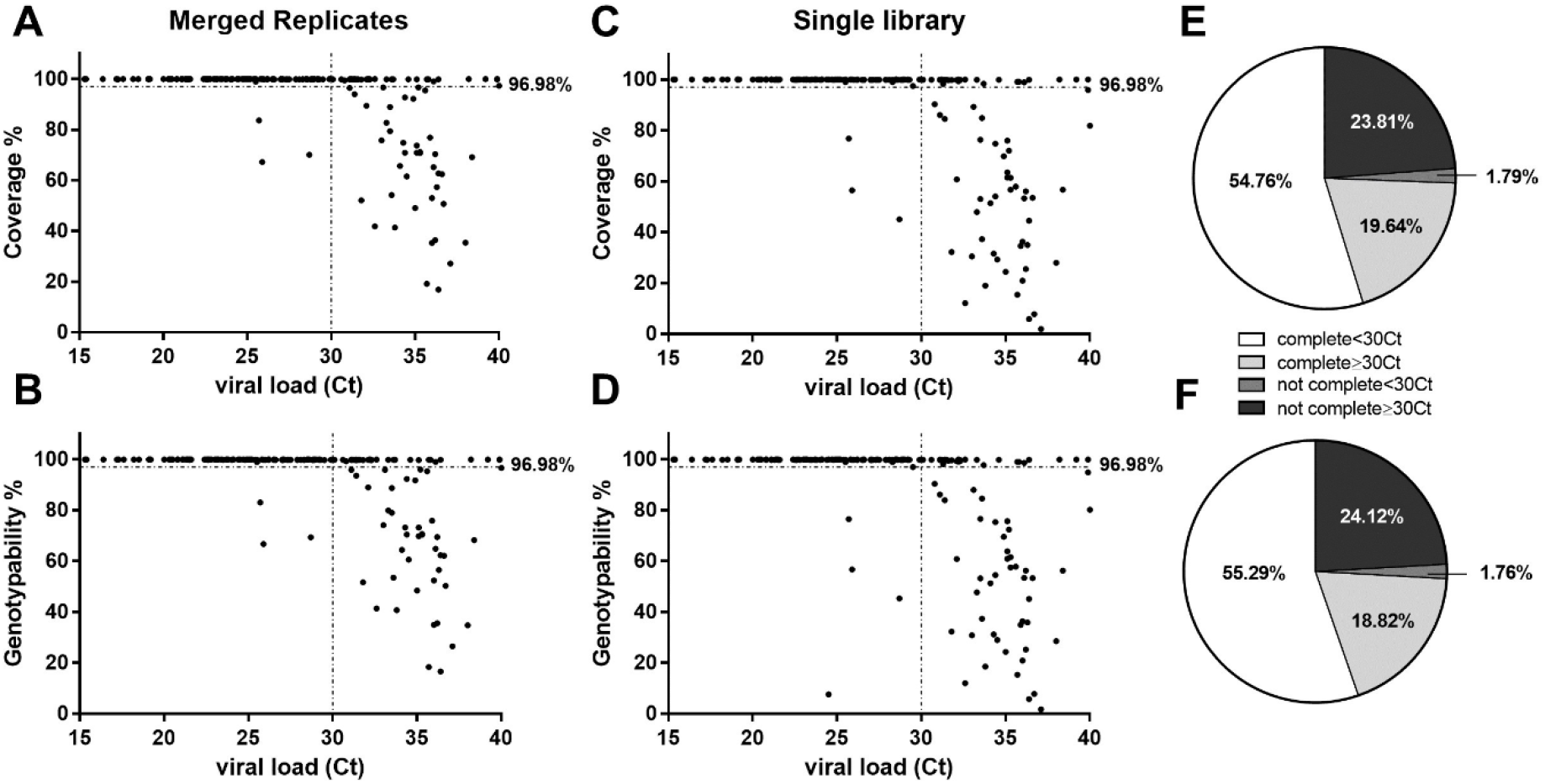
SARS-CoV-2 sequencing in a cohort of clinical samples with wide range of viral titers. **(A-C)** Percentage of genome coverage and **(B-D)** genotypability for each sample (N = 170) considering a single replicate (selected randomly) or after merging two sequencing replicates. The pie charts show the fraction of the complete SARS-CoV-2 (>96.98%) genome in terms of **(E)** coverage or **(F)** genotypability for samples with Ct < or ≥ 30.

### Impact of genome reconstruction accuracy on the evaluation of a potential re-infection case

The identification of SARS-CoV-2 genetic variants at different time points can reveal whether recurrent infections are relapses caused by the same strain or independent infections with a different strain. We therefore evaluated our optimized workflow in a case-study of relapse/re-infection involving a 48-year-old female patient who was hospitalized with mild COVID-19 symptoms following a positive nasopharyngeal swab on 4/3/2020, discharged with no symptoms on 11/3/2020 followed by two consecutive negative swab tests, but readmitted with mild COVID-19 symptoms 12 days later. During the second hospital stay, the nasopharyngeal swab test results fluctuated, and the patient was finally discharged on 21/4/2020 with no symptoms, and two consecutive negative molecular tests. Three swab samples (one from the first and two from the second hospitalization period) were sequenced to identify the viral strain responsible for infection (**Table 1**). All samples were sequenced in duplicate or quadruplicate (**Table S14**), and consensus variants were called in order to identify the viral strains. Depending on the replicate, some consensus variants identified in the first hospitalization period were missing or could not be genotyped in the second hospitalization period, leading to the hypothesis that different strains could be responsible for each infection (**Table 1**). In contrast, when merging sequencing replicates, the same variants were identified in all three samples (**Table 1**) and a very high-frequency (99.95%) false-positive variant could be identified at position 12890 (**Table S13**). Based on this analysis, we concluded that the same viral strain was responsible of both the first and second infection, and that the latter should therefore not be classed as a re-infection.

**Table 1.**
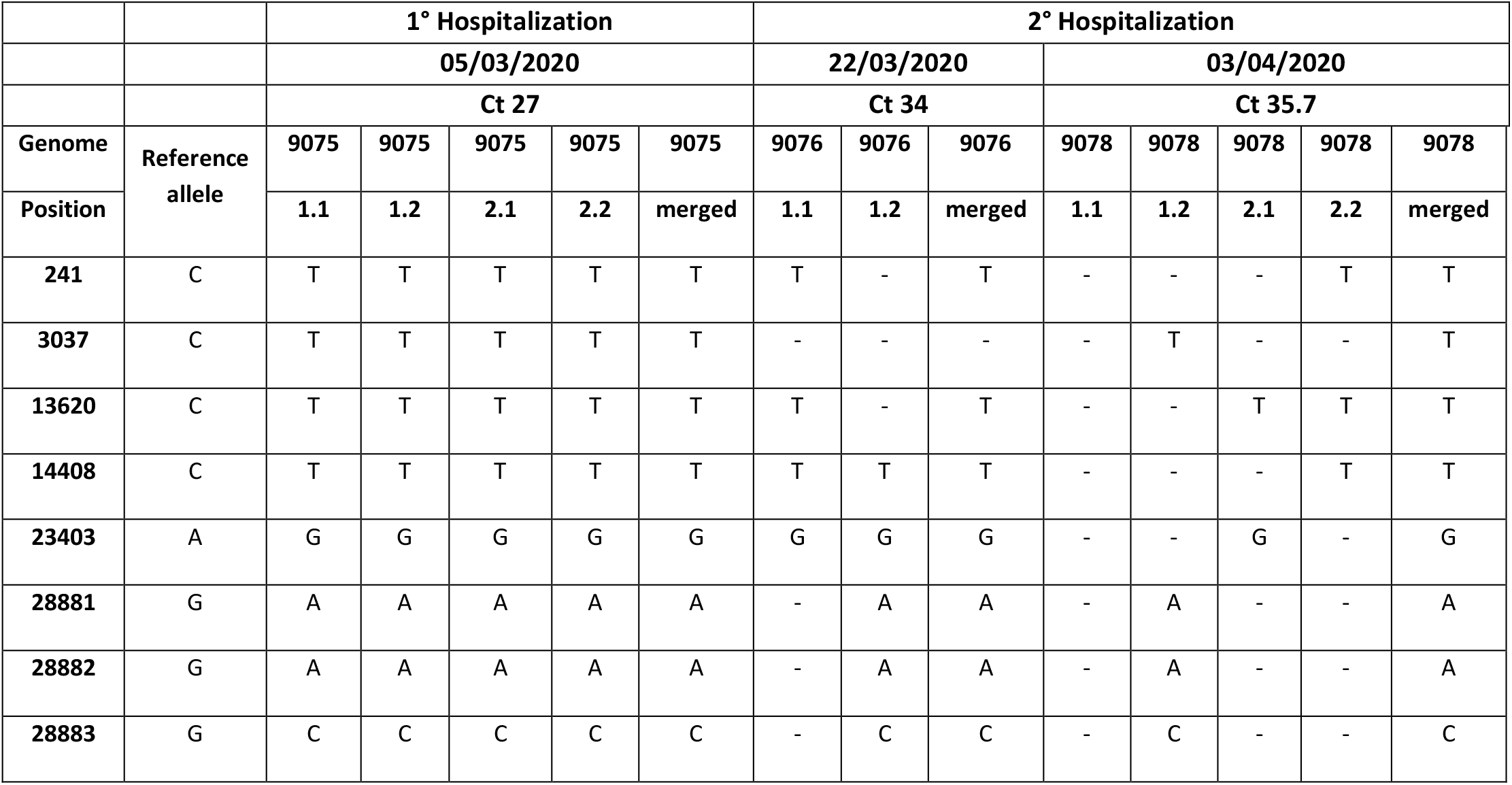
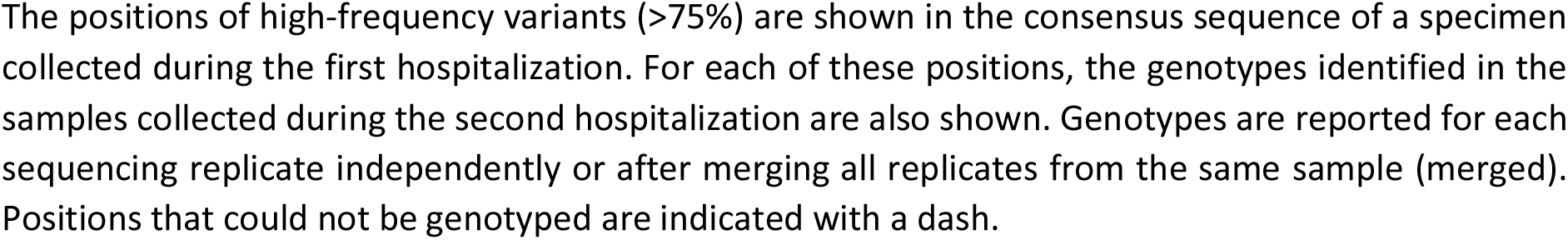
High-frequency variants identified in the COVID-19 relapse case study.

|  |  | 1° Hospitalization |  |  |  |  | 2° Hospitalization |  |  |  |  |  |  |  |
| --- | --- | --- | --- | --- | --- | --- | --- | --- | --- | --- | --- | --- | --- | --- |
|  |  | 05/03/2020 |  |  |  |  | 22/03/2020 |  |  | 03/04/2020 |  |  |  |  |
|  |  | Ct 27 |  |  |  |  | Ct 34 |  |  | Ct 35.7 |  |  |  |  |
| Genome | Reference allele | 9075 | 9075 | 9075 | 9075 | 9075 | 9076 | 9076 | 9076 | 9078 | 9078 | 9078 | 9078 | 9078 |
| Position |  | 1.1 | 1.2 | 2.1 | 2.2 | merged | 1.1 | 1.2 | merged | 1.1 | 1.2 | 2.1 | 2.2 | merged |
| 241 | C | T | T | T | T | T | T | - | T | - | - | - | T | T |
| 3037 | C | T | T | T | T | T | - | - | - | - | T | - | - | T |
| 13620 | C | T | T | T | T | T | T | - | T | - | - | T | T | T |
| 14408 | C | T | T | T | T | T | T | T | T | - | - | - | T | T |
| 23403 | A | G | G | G | G | G | G | G | G | - | - | G | - | G |
| 28881 | G | A | A | A | A | A | - | A | A | - | A | - | - | A |
| 28882 | G | A | A | A | A | A | - | A | A | - | A | - | - | A |
| 28883 | G | C | C | C | C | C | - | C | C | - | C | - | - | C |

The positions of high-frequency variants (>75%) are shown in the consensus sequence of a specimen collected during the first hospitalization. For each of these positions, the genotypes identified in the samples collected during the second hospitalization are also shown. Genotypes are reported for each sequencing replicate independently or after merging all replicates from the same sample (merged). Positions that could not be genotyped are indicated with a dash.

## DISCUSSION

### Protocol optimization for simplicity, flexibility, throughput and cost-efficiency

Amplicon-based sequencing (originally called PrimalSeq) is the most sensitive and widely-used protocol for SARS-CoV-2 whole-genome analysis from clinical isolates, but its disadvantages include uneven amplicon coverage and poor accuracy when the viral load is low [23]. We addressed these limits by improving the accuracy and completeness of sequencing, as well as the cost-efficiency and throughput, thus achieving the highly reliable analysis of SARS-CoV-2 genomes. This benchmarking analysis established a robust workflow, ACoRE, that allowed the complete and accurate characterization of SARS-CoV-2 genomes in 170 clinical samples, including a subset (42%) with very low viral titers (Ct ≥ 30). We were also able to properly categories an infection-relapse case study.

The protocol optimized by the ARTIC Network for SARS-CoV-2 genome sequencing utilizes a tiling primer scheme generating 400-bp viral amplicons for adaptor ligation and 250PE sequencing [33]. This limits the sequencing options on Illumina platforms because this read type is compatible only with the MiSeq v2 chemistry and NovaSeq6000 SP flow cells. To increase flexibility, we used the NexteraFlex kit to prepare amplicon libraries with shorter inserts (170–200 bp) suitable for 150PE sequencing without loss of performance. This also confers the ability to pool up to 384 samples in a single run using unique dual indexes, reducing costs from €80 per sample to €3.5 on the NovaSeq6000 with S1 flow cell or €12 on the NextSeq500 with HighOutput flow cell. Even shorter sequencing reads (100PE) resulted in shorter overlap of paired ends, reducing the number of sequencing fragments required per sample and translating to even lower costs of €3 per sample. Because the NexteraFlex method does not require the quantification of starting amplicons or final sequencing libraries, this further reduces costs and processing time. Further savings could potentially be achieved by using half the volume of tagmentase reagent, but testing is required to ensure that accuracy and coverage is maintained. The generation of amplification replicates from a single starting cDNA (instead of multiple cDNAs, as recommended by the original protocol[23]) would also save time and costs, while preserving the sample for additional tests. The fragmented amplicon approach and other adjustments therefore improved protocol simplicity, flexibility, multiplexing and economy, allowing the cost-effective and timely processing of larger cohorts of samples by ACoRE.

### Sequencing multiple replicates to increase accuracy and completeness

Clinical specimens with low viral loads reduce the accuracy of variant calling and the completeness of genome reconstruction, both of which are inversely correlated with the quality and quantity of starting material[23,30,43]. Current guidelines for viral genotyping recommend a lower limit of 1000 virus copies per reaction [23,43] but this would rule out a large proportion of clinical samples, including ∼53% of the samples in our cohort. A Ct value of ∼25 was identified as the median for virus detection in symptomatic patients, with a consistent proportion of samples (15–25%) falling above Ct 30 [25,46]. Low viral loads are often found in patients with prolonged COVID-19 infection [47–49], and five of six reported cases of potential re-infection involved samples with Ct values >30 [50], but whole-genome sequencing is nevertheless recommended to differentiate between relapse and new infections caused by a different SARS-CoV-2 variants [50,51]. The ability to sequence SARS-CoV-2 genomes in low-titer samples is therefore necessary to track infections and correlate different strains with disease communicability, manifestation and severity.

Increasing the depth of sequencing has been proposed as a strategy to achieve complete genome reconstruction in low-titer samples, but this does not overcome limitations caused by missing amplicons [43]. Similarly, improvement in ARTIC primer design and compatibility (currently version 3) can also ameliorate genome coverage, but again cannot make up for missing amplicons [24,30]. We found that only a few specific amplicons were reproducibly suboptimal (64, 70 and 91) whereas most showed coverage variations limited to particular samples or replicates. We therefore merged the sequencing data from two or more replicates as a simple solution to enhance coverage and genotypability, achieving a more homogeneous representation of the viral genome and rescuing the suboptimal samples. The random amplification observed in low-titer samples most likely reflects the low sample complexity rather than poor assay sensitivity or performance. Accordingly, the sampled RNA and corresponding cDNA fragments before amplification are unlikely to represent the complete genome based on our observation that the coverage achieved by sequencing two amplification replicates (each from 5 µL of cDNA) was similar to that achieved with a single amplification starting from double the amount of cDNA (10 µL). Therefore, to optimize genome reconstruction, a single large cDNA batch should be amplified in several parallel reactions, using as much sample volume as possible to increase complexity. The multiple PCR products can then be pooled before library preparation and sequenced as a single sample to avoid increasing costs.

As well as improving coverage and genotypability, at least two amplification reactions must be analyzed to achieve accurate variant calling (SNVs and iSNVs). It is well established that the analysis of viral iSNVs down to 3% frequency requires the generation of multiple replicates to distinguish true-positive iSNVs from low-frequency PCR or sequencing errors [23]. In contrast, the generation of consensus sequences for the analysis of SNVs in epidemiological studies requires the identification of the most-frequent nucleotide at each position and is typically based on single replicates [12,45]. However, we discovered that consensus sequences also contain frequent SNV errors (>12% in our cohort) and the comparison of technical replicates is required to ensure accuracy. This was not confined to low-titer samples (Ct > 30) but also included some samples with moderate viral loads (Ct = 25–30) potentially leading to the submission of inaccurate consensus sequences to public repositories such as GISAID. These false-positive variants probably arose due to PCR errors because they were not found in other amplification replicates (either from the same or different cDNA). However, studies reporting SARS-CoV-2 consensus sequences thus far have not included the analysis of technical replicates, even in the case of low-titer samples (Ct > 30)[26,52]. The accuracy of SARS-CoV-2 consensus sequences deposited in GSAID has been called into question for documented sequences with putative errors or a significant number of variants in one particular submission (singletons) [35] and the use of stringent filters and bioinformatic tools has been proposed as a solution [52,53]. Instead, with ACoRE we propose the use of replicates as a simple experimental solution to avoid the generation of incorrect consensus sequences prior to database submission.

### The assessment of re-infections

Reconstruction of highly accurate sequences from sub-optimal samples was crucial to identify the correct viral strain responsible of a second hospitalization case, that was hypothesized to be a re-infection. A standard workflow would have missed or included incorrect variants in support of such hypothesis, while ACoRE properly recognized that the different time-point samples contained the same viral strain.

Another interesting example, that would certainly benefit of ACoRE, comes from a publication that reported the first individual in North America to have symptomatic reinfection with SARS-CoV-2 [26], for whom “…genomic analysis of SARS-CoV-2 showed genetically significant differences between each variant associated with each instance of infection…” suggesting that “…the patient was infected by SARS-CoV-2 on two separate occasions by a genetically distinct virus…” [45]. The viral load of the swab samples analyzed in that study was very low (Ct > 35) based on 14–22 PCR cycles-protocol without amplification replicates, therefore potential false-positive variants and/or regions with low genotypability may have influenced the results. We reanalyzed the data and noted that two of the four variants specifically associated with the first infection had insufficient sequencing coverage to achieve confident variant calling in the sample from the second infection (**Table S15**). In particular, our bioinformatic pipeline revealed that position 539 was covered by only five reads, thus a genotype could not be properly called; while variant 16741G→T (supported by 10 reads) was only just above the genotypability threshold of 8 (**Table S15**). These positions were genotyped using the bioinformatic pipeline utilized by the authors because the limit was set to five reads. Furthermore, variant 4113C→T showed frequency of 67.82% in the first infection, suggesting that two viral strains were already present: a predominant strain carrying the identified variant and a less-abundant strain lacking the variant that became prevalent in the second infection (**Table S15**). However, the absence of replicate analysis makes it impossible to confirm this hypothesis. Similarly, although the final variant (7921A→G) was abundant, the absence of replication makes it impossible to rule out the possibility of an amplification error, as frequently observed in our low-titer samples. These questions could be resolved by sequencing two technical replicates rather than analyzing data from one sequencing library using two different pipelines (as reported by the authors). The conclusions put forward by the authors therefore appear to be only weakly supported by the raw data, but would nevertheless have a major impact on future research by highlighting the possibility of re-infection and thus possibly questioning the efficacy of vaccines. The analysis of such critical samples would greatly benefit from the use of technical replicates, and robust evaluation is particularly important due to the ramifications of the conclusions for the global research and biomedical communities.

## CONCLUSIONS

We have optimized ACoRE, a workflow for SARS-COV-2 sequencing to improve flexibility and throughout, thus reducing assay time and costs and facilitating the robust analysis of suboptimal samples that would normally be excluded from sequencing even if they are central and irreplaceable specimens. The sequencing of such low-titer samples without replication risks the generation of consensus sequences containing false-positive SNVs and iSNVs, but we found that the inclusion of technical replicates improves both the accuracy and completeness of viral genome analysis. This reduces the risk of generating inaccurate and incomplete genomic sequences, favoring the submission of robust sequences to public databases and enhancing the downstream analysis of SARS-CoV-2 genotyping data.

## METHODS

### Clinical samples

178 Nasopharyngeal swabs (eSwab, Copan, Italy) were obtained from 172 COVID-19 patients diagnosed at the Department of Infectious, Tropical Diseases and Microbiology of the IRCCS Sacro Cuore Don Calabria Hospital, qualified for SARS-CoV-2 molecular diagnosis by the regional reference laboratory (Department of Microbiology, University Hospital of Padua). After collection, swabs were stored at 4 °C for a maximum of 48 h and analysed by the routine-used molecular diagnostic method (RT-qPCR as indicated in the following paragraph). The remaining quantity of swab was then aliquoted and preserved at –80 °C. The study was approved by the competent Ethical Committee for Clinical Research of Verona and Rovigo Provinces (Prot N° 39528/2020).

### RNA extraction and RT-qPCR analysis

The routine RT-qPCR protocol was based on a recommended test (emergency use authorization) standardized according to I asked what I’m reading WHO guidelines. Briefly, RNA was extracted from 200μL of swabs using the automated Microlab Nimbus workstation (Hamilton, Reno, NV, USA) coupled to a Kingfisher Presto system (Thermo Fisher Scientific, Waltham, MA, USA). We also used a MagnaMax Viral/Pathogen extraction kit (Thermo Fisher Scientific) according to the manufacturer’s instructions. RT-qPCR was carried out using the CDC 2019-nCoV rRT-PCR Diagnostic Panel assay and protocol [54], targeting the nucleocapsid protein gene regions N1 and N2 (with the human RNAse P gene as the internal control) on a CFX96 Touch system (Bio-Rad Laboratories, Milan, Italy) with white plates. The amplification cycle threshold (Ct) was determined using CFX Maestro (Bio-Rad Laboratories), setting a baseline threshold at 200 relative fluorescence units (RFU). A standard curve from 5 to 500 genome copies per reaction was performed with serial dilution of the CDC control plasmid containing the complete nucleocapsid gene of SARS-CoV-2 (**Table S1**).

### Reverse transcription and amplification of the SARS-CoV-2 genome

Samples with Ct values of 15–18 were diluted 10-fold as suggested by the ARTIC Network [27]. RNA from swab samples (5 µL) was first incubated with 1 µL of 60 μM Random Primer Mix (New England Biolabs, Ipswich, MA, USA) and 1 µL of 10 mM dNTPs (New England Biolabs) at 65 °C for 5 min followed by 1 min on ice to anneal the primers. We then added 4 µL of 5× SSIV buffer, 1 µL of 100 mM DTT, 1 µL of 40 U/μL RNaseOUT, 1 µL of 200 U/μL SSIV enzyme (Thermo Fisher Scientific) and 6 µL nuclease-free water (total reaction volume = 20 µL) and heated the reaction to 23 °C for 10 min, 52 °C for 10 min and 80 °C for 10 min. We generated two or three cDNAs from each sample (depending on the experiment), each of which was amplified 2–3 times using the ARTIC protocol. In each case, we mixed 2.5 or 5 µL cDNA (depending on the experiment) with 3.7 µL of 10 µM primer pools A and B from the ARTIC nCoV-2019 V3 panel (IDT, Coralville, IA, USA), 12.5 µL Q5 high-fidelity DNA polymerase 2× (New England Biolabs) for each of the primer pools, and nuclease-free water to a final volume of 25 µL. The reaction was heated to 98 °C for 30 s, followed by 25 cycles (sample Ct ≤ 21) or 35 cycles (sample Ct > 21) of 98 °C for 15 s and 65 °C for 5 min. The PCR products were then combined in a single tube, cleaned up using 1× AMPure XP beads (Beckman Coulter, Brea, CA, USA) and eluted in 15 µL of water. The resulting amplicons were analyzed on the 4150 TapeStation System (Agilent Technologies, Santa Clara, CA, USA) and quantified using the Qubit dsDNA HS Assay kit (Thermo Fisher Scientific).

### Full-length amplicon sequencing

Libraries were prepared from 50 ng of virus amplicons using the KAPA Hyper prep kit and unique dual-indexed adapters (5 µL of a 15 µM stock) according to the supplier’s protocol (Roche, Basel, Switzerland). The post-ligation products were cleaned up using 0.8× AMPure XP beads followed by library amplification (six cycles) with the KAPA Library Amplification Primer Mix (Roche). After a clean-up with 1× AMPure XP beads, the libraries were analyzed on the 4150 TapeStation System (average size 526–573 bp) and quantified using the Qubit dsDNA BR Assay kit (Thermo Fisher Scientific). Barcoded libraries were pooled at equimolar concentrations and sequenced on the MiSeq platform (Illumina, San Diego, CA, USA) with Miseq Reagent kit v2 to generate 250-bp paired-end (250PE) reads.

### Fragmented amplicon sequencing

Libraries were prepared from 10 µL of purified viral amplicons using the NexteraFlex kit (Illumina) according to the manufacturer’s recommendations, and combinatorial dual indexes were added in six cycles of PCR. We cleaned up 10-µL aliquots of each amplified library using a 1:1 ratio of sample purification beads (Illumina) and eluted the purified library in 20 µL of resuspension buffer (Illumina). The resulting libraries were analyzed on the 4150 TapeStation System (average size 335–369 bp), pooled and quantified using the Qubit dsDNA BR Assay kit. The libraries were sequenced on a Novaseq 6000 device (Illumina) using an SP flow cell to generate 100-bp paired-end (100PE) reads, or on a NextSeq500 (Illumina) to generate 150-bp paired-end (150PE) reads.

### Data filtering and reference genome alignment

Full-length amplicon sequencing data were randomly downsampled using *seqtk sample v1*.*3* (https://github.com/lh3/seqtk). To compare sequencing data from the full-length and fragmented amplicons, KAPA library reads were downsampled at the same mean mapped coverage as the corresponding NexteraFlex replicates using *sambamba v0*.*6*.*7* [55]. To simulate sequencing using 100PE reads, data from the fragmented amplicon libraries were trimmed using a custom script. All sequencing datasets were trimmed for quality and adapters were removed using *Trimmomatic v0*.*39* [56] with the following parameters: *ILLUMINACLIP:adapters_file:2:30:10 LEADING:5 TRAILING:5 SLIDINGWINDOW:4:20*. Filtered reads were aligned to the SARS-CoV-2 reference genome (GenBank ID: *MN908947*.*3*) using *BWA MEM v0*.*7*.*17* [57] with default parameters and the relative alignment file was converted to a binary alignment map (BAM) file using *SAMtools v1*.*9* [58]. For the fragmented libraries, duplicate reads were identified and discarded using *Picard v2*.*21*.*1* (http://broadinstitute.github.io/picard). Subsequently, *iVar v1*.*2*.*2 trim* [23] was used to remove ARTIC v3 primer sequences from the BAM files. For the fragmented libraries, the -*e* parameter was used to include reads without primers. Finally, overlapping portions of reads were clipped using *fgbio ClipBam v1*.*1*.*0* (https://github.com/fulcrumgenomics/fgbio) with the following parameters: *--clip-overlapping-reads -c Hard* to avoid counting multiple reads representing the same fragment. Coverage and genotypability statistics were calculated from the BAM files using *bedtools genomecov v2*.*19*.*1* [59] and *GATK CallableLoci v3*.*8* [60], respectively. Raw genomic sequencing data were deposited in NCBI GenBank (BioProject no PRJNA690890).

### Consensus variant calling and generation of the consensus sequence

A pileup was calculated for each position in the BAM file of each replicate using the *SAMtools v1*.*9 mpileup* option with parameters *-aa -A -d 0 -Q 0*. The resulting files were used as input for *iVar consensus v1*.*2*.*2* [23] to generate consensus sequences, considering those positions covered by at least three reads (parameters: *-t* 0 -m 3). The most abundant nucleotide for each position was reported in the consensus sequence, whereas positions covered by fewer than three reads or reporting an equal proportion of nucleotides were represented by the ambiguous character N.

To call variants present in the consensus sequences (consensus variants), sequences were aligned to the SARS-CoV-2 reference genome using *Minimap v2*.*17* [61] and the alignment file was converted to the BAM format using *SAMtools v1*.*9*. Consensus variants were then called using *bcftools call v1*.*10*.*2* [58] with the following parameters: *--ploidy 1 -A -m -P 0*.*05 -M -Oz*.

Final consensus sequences from the cohort of 170 samples and the relapse case were called after merging sequencing data for each individual replicate. False-positive variants in the consensus sequence were identified manually by comparing the presence of discordant iSNVs at the same genomic position between replicates of the same sample and considering only positions genotyped in both replicates. False-positive variants were removed from consensus sequences and replaced with the reference allele.

### iSNV variant calling

Alignment BAM files were used to call iSNVs present in each replicate with a minimum minor allele frequency (MAF) threshold of 3%. Joint variant calling of the 30 entire amplicon libraries, and between replicates of the same sample for fragmented amplicon libraries, was achieved by generating a pileup using S*AMtools mpileup v1*.*9* [58] with the following parameters: *-A -d 600000 -B -Q 0*. The output file was used to detect iSNVs with *VarScan mpileup2cns v2*.*3*.*9* [62] and the following parameters: *--min-var-freq 0*.*03 --min-avg-qual 20*.

For each sample, inter-replicate discordant variants were identified by iSNV variant calling after merging sequencing data from all replicates, considering only genotyped positions. A discordant variant was defined as a variant called in one replicate, whereas the same position in the other replicate reported the reference allele.

### Calculation of the concordance rate

The concordance rate (R_c_) between replicates samples was calculated as follows:

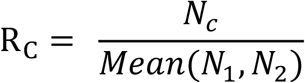

*Nc* represents (i) the number of shared variants (consensus variants or iSNVs) excluding positions that could not be genotyped in at least one replicate, or (ii) the number of shared genotypable bases, excluding positions marked N in at least one replicate, or (iii) the number of shared amplicons with coverage higher than three reads in all replicates. *N1* and *N2* represent the total number of iSNVs, consensus variants, genotypable bases or covered amplicons detected in each of the two samples in the analysis. R_c_ was calculated by comparing couples of replicates generated from the same cDNA (intra-cDNA concordance) and triplets of replicates generated from different cDNAs (inter-cDNA concordance) as shown in **Table S2**.

### Statistical analysis

The non-parametric Wilcoxon signed rank test and the Mann Whitney U-test were used to compare matched pairs and non-matched data, respectively. The non-parametric Friedman test was used to compare multiple paired groups. Significance of pairing was confirmed by calculating Spearman’s rho. We used GraphPad Prism 6.0 (GraphPad Software, San Diego, CA, USA) for all statistical analysis, with a significance threshold of p < 0.05.

## Data Availability

all fastq data have been upload to the repository

http://ddlab.sci.univr.it/files/SARS-CoV2_Paper/SARS-CoV2_Paper.tar

## DECLARATIONS

### Ethics approval and consent to participate

The study was approved by the competent Ethical Committee for Clinical Research of Verona and Rovigo Provinces (Prot N° 39528/2020)

### Consent for publication

Not applicable.

### Availability of data and materials

The raw reads dataset supporting the conclusions of this article is available at the NCBI SRA repository under BioProject ID PRJNA690890.

### Competing interests

The authors declare that they have no competing interests

### Funding

The work performed at IRCCS Sacro Cuore Don Calabria Hospital was supported by the Italian Ministry of Health “Fondi Ricerca corrente—L1P5”.

## Authors’ contributions

The study was conceived and coordinated by MD and MR. The samples and RNA extraction and RT-qPCR analysis were provided and performed by CP, AM, EP, MD, SS, ZB. Reverse transcription, amplification of the SARS-CoV-2 genome, library preparation and sequencing were performed by CB, CDE, VG, EM, MP, ES, BG. The data filtering and reference genome alignment, the consensus variant calling and generation of the consensus sequence, the iSNV variant calling, the calculation of the concordance rate and the statistical analysis were performed by LM, GL, SM, DL, BI, MG. The manuscript was written by MR with input from all co-authors. All authors read and approved the final manuscript

## Acknowledgements

We gratefully acknowledge the Centro Piattaforme Tecnologiche (CPT) for granting access to the genomic facility of University of Verona for sequencing on a MiSeq and NextSeq500 Illumina platform, and Dr. Richard M Twyman (www.twymanrm.com) for editing the manuscript text.

## Notes

### Competing Interest Statement

The authors have declared no competing interest.

### Author Declarations

The study was approved by the competent Ethical Committee for Clinical Research of Verona and Rovigo Provinces (Prot N 39528/2020)

