## Supplemental figures for "ACoRE: Accurate SARS-CoV-2 genome reconstruction for the characterization of intra-host and inter-host viral diversity in clinical samples and for the evaluation of re-infections"

**Figure S1. Sequencing coverage of the 30 intra-cDNA and inter-cDNA replicates.** Sequencing coverage of the 98 ARTIC V3 panel amplicons for the 30 intra-cDNA and inter-cDNA replicates generated from five COVID-19-positive swab samples. Green bars represent the amplicons generated using the ARTIC original primer set, and orange bars show the amplicons generated using the alternative V3 primers.

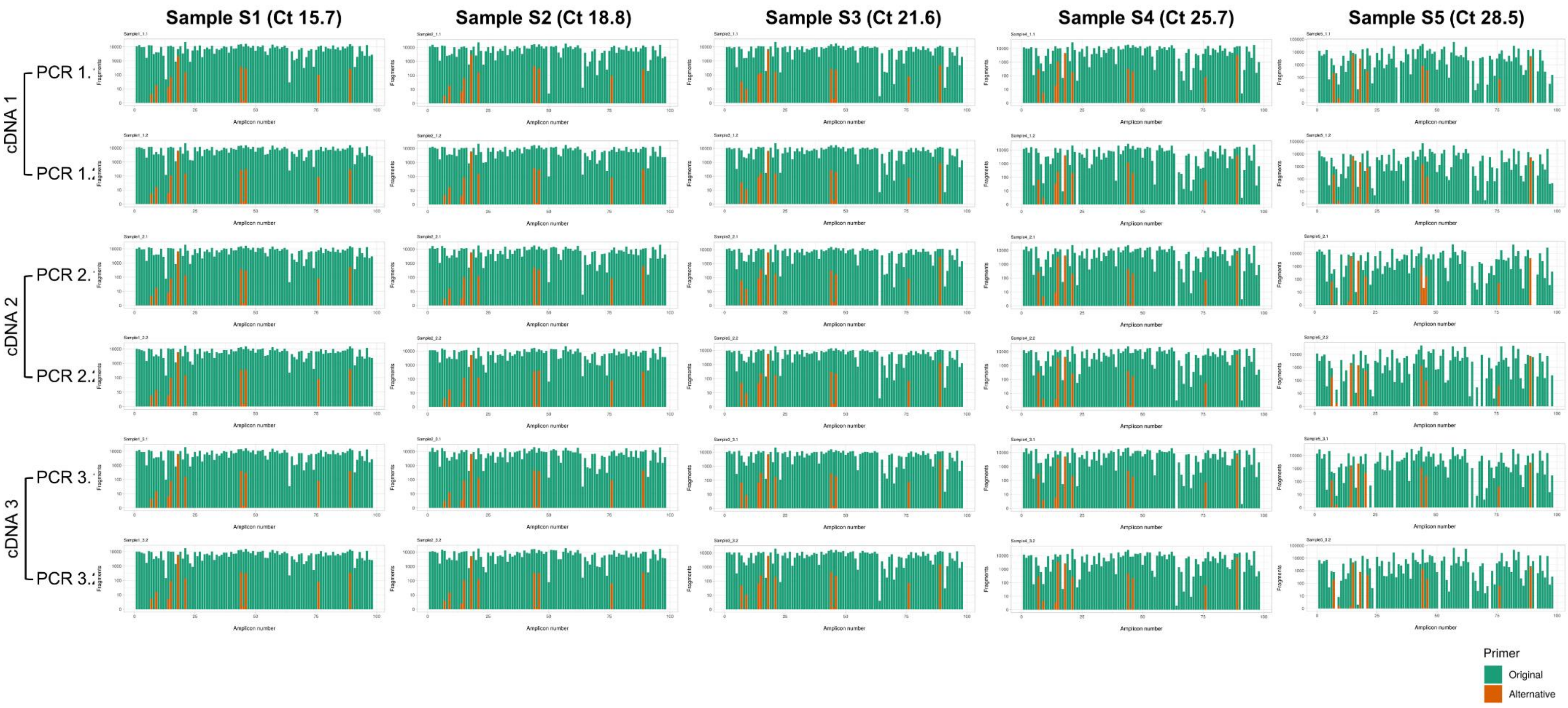

**Figure S2. Coverage and genotypability are similar when amplifying higher cDNA volumes or pooling distinct replicates. (A)** Ct frequency distribution of the 20 COVID-19-positive samples analyzed in (B) and (C). Ct values are rounded to the closest integer number. **(B)** Coverage and **(C)** genotypability when analyzing a single replicate generated from 10 µL cDNA or after merging the sequencing results of two replicates generated from 5 µL cDNA.

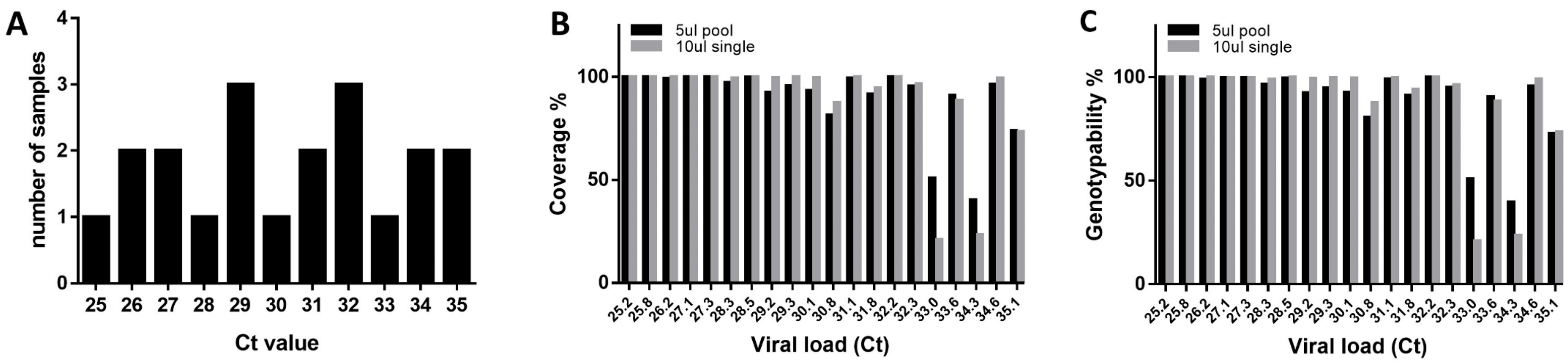

**Figure S3. Distribution of Ct values across a cohort of 170 COVID-19 swab samples.** The graph shows the frequency of COVID-19 swab samples showing a certain Ct (determined by RT-qPCR) in the cohort analyzed in Figure 5. Ct values are rounded to the closest integer number.

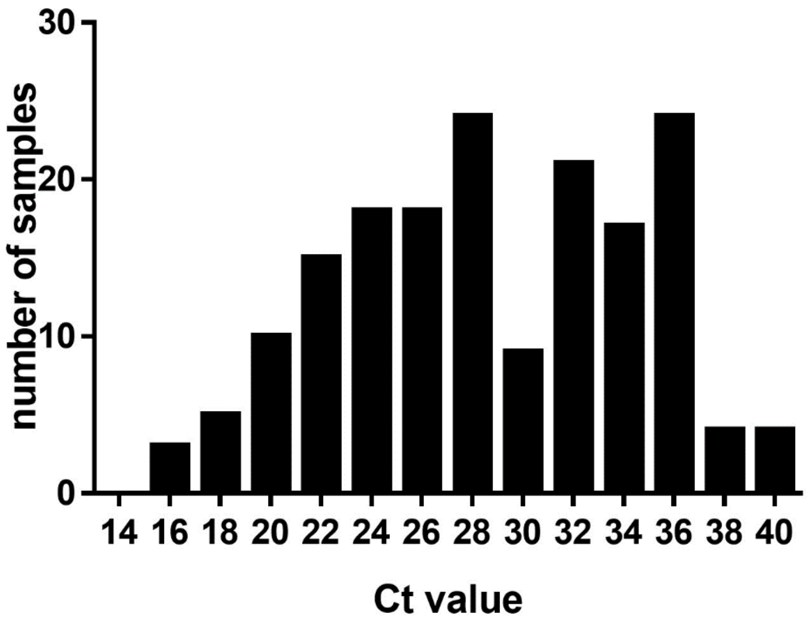
